## Supplementary Information for "Estimating the potential impact and diagnostic requirements for SARS-CoV-2 test-and-treat programs"

**Supplemental information**

### Estimating the potential impact and diagnostic requirements for SARS-CoV-2 test-and-treat programs in low-and-middle-income countries

Alvin X. Han[1], Emma Hannay[2], Sergio Carmona[2], Bill Rodriguez[2], Brooke E. Nichols[1,2,3,†], Colin A. Russell[1,3,†]

[1]Department of Medical Microbiology & Infection Prevention, Amsterdam University Medical Center, University of Amsterdam, Amsterdam, The Netherlands
[2]Foundation for Innovative New Diagnostics (FIND), Geneva, Switzerland
[3]Department of Global Health, School of Public Health, Boston University, Boston, MA, USA
†Contributed equally

**Supplementary Text**

**Additional details of the Propelling Action for Test And Treat (PATAT) model**

*Population demography*

Using input demographic data which includes information such as population age and sex distribution, household composition, employment and schooling rates, PATAT generates a population of individuals who are linked by a series of underlying contact network settings where transmission may occur. These contact network settings include households, schools, workplaces, regular mass gatherings (i.e. church) as well as random community contacts.

*Household*

PATAT randomly generates a Poisson distribution of household sizes based on the given mean household size. A reference individual (e.g. head of the household) above an assumed prime adult age (e.g. years) is first randomly assigned to each household. To account for multigenerational households, the remaining household members are then randomly sampled multinomially by the input age distribution of households. Although PATAT does not explicitly model the geolocation of agents, households are ordered to implicitly approximate neighbourhood proximity. Herein, households are assigned with a numerical identifier. The smaller the difference between the household numerical identifiers, the nearer the households were assumed to be by location.

*Schools*

PATAT distinguishes between elementary and secondary schools. For each education level, schooling children are randomly sampled from the population based on given enrolment rates and gender parity. Class sizes are then randomly drawn from a Poisson distribution based on the input mean class size while constrained by the number of schooling children attending the same grade (i.e. age; a class include only students studying the same grade). Schools are created by random allotment of classes such that (1) all schools will have equitable distributions of classes of all grades for the given education level and (2) the total number of students approximately equals to the expected school size. Classes are then populated by schooling agents such that (1) agents of proximally ordered households will tend to attend the same school and (2) children of the same grade (age) from identical households will not be assigned to the same class even though they may attend the same school. School teachers are then randomly drawn from the employed prime adult population based on the input teacher-to-student ratio and are assumed to have contact with each other during school days. Each class is randomly assigned to one teacher.

*Workplaces*

PATAT generates both formal and informal workplace contact networks based on separate employment rates. Youth (15-19 years) employment is also considered in the potential workforce. The distinction between formal and informal settings is made as mean employee contact rates likely differ between them. Furthermore, workplace distribution of Ag-RDTs for community testing is assumed to be feasible for formal employment entities only. Unlike schools, PATAT does not explicitly model for workplaces but sets up contact matrices between employed individuals who would be in regular contact at work. As such, different number of formal and informal mean number of work contacts must be provided by the user and sizes of workplace contact network are randomly drawn from a Poisson distribution. An employed agent would only be associated with one workplace contact network.

*Mass gatherings (Religious gathering)*

High-density mass gatherings are considered in the model in the form of contacts among church congregations. The size of a church is assumed to follow a Normal distribution with user's given mean and variance. PATAT assumes that all members of a household will visit a church together every Sunday. Other than close contacts with each other, each household member would also have a random number of close contacts from other households that attend the same church. This random contact number is drawn from a Gamma distribution with user's given shape and scale parameters. Churches are also ordered such that proximally ordered households in the same neighbourhood would visit the same church.

*Random community*

PATAT assumes that every agent within a given age range would have a random number of contacts with the community daily, drawn from a Poisson distribution with a mean defined by the user.

*Model Validation*

To validate our model, we used the estimated percentage population that would test positive for COVID-19 infection, regardless of whether they reported that they were experiencing symptoms, during the spread of Omicron BA.5 (effective reproduction number ($R_e$) = ~1.5) and XBB.1.5 ($R_e$ = ~1.2) subvariants in the UK, including England, Wales and Scotland (https://www.ons.gov.uk/peoplepopulationandcommunity/healthandsocialcare/conditionsanddiseases/bulletins/coronaviruscovid19infectionsurveypilot/24march2023). These estimates reflect the community prevalence of SARS-CoV-2 in the UK based on testing results from random community supervised self-swabbing RT-PCR-based surveillance collected across the country[1] (~300,000 swab tests per month; https://www.ons.gov.uk/peoplepopulationandcommunity/healthandsocialcare/conditionsandd

iseases/methodologies/coronaviruscovid19infectionsurveyqmi for more details). For the same variant $R_e$ values, the UK estimates fitted well within the 95% bootstrap confidence interval of the proportion of infectious individuals, regardless if they presented symptoms, that would test positive over time in our PATAT simulations.

#### Supplementary Figures

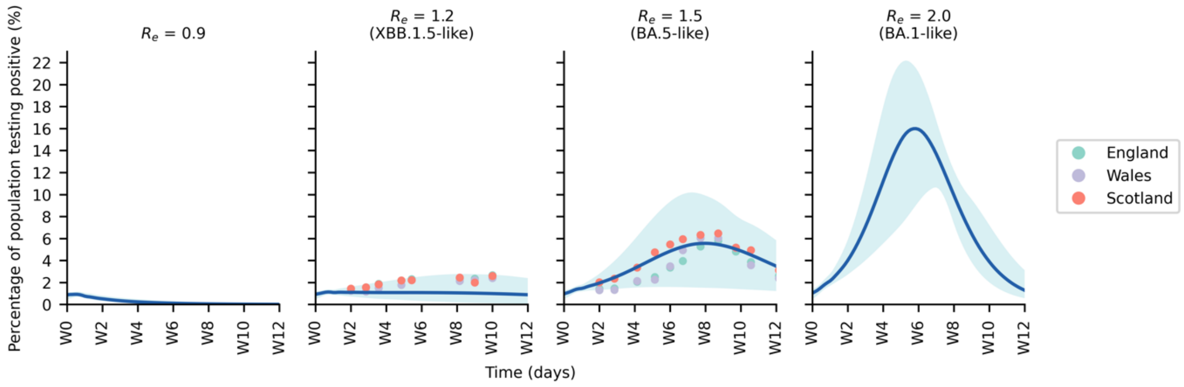


**Figure S1**: **Model validation – Comparing simulated prevalence against estimated UK prevalence**. The blue line shows the average simulated percentage of the population that would test positive for COVID-19 each day (i.e. percentage of individuals that were productively infectious, regardless if they were symptomatic, and would test positive for SARS-CoV-2) for different effective reproduction number ($R_e$). The blue shaded area shows the range of simulated percentages across all simulations for all four simulated countries (i.e. Zambia, Brazil, Georgia and the Netherlands), under all testing rates and vaccination coverage, with *no* test-and-treat programs. The scatter points in subplots of $R_e$ = 1.2 and $R_e$ = 1.5 show the estimated community prevalence data from different countries (i.e. England, Wales and Scotland) in the UK (https://www.ons.gov.uk/peoplepopulationandcommunity/healthandsocialcare/conditionsanddiseases/bulletins/coronaviruscovid19infectionsurveypilot/24march2023) during the spread of BA.5 ($R_e$~1.5) and XBB.1.5 ($R_e$~1.2). The UK prevalence estimates were based on testing results from random community supervised self-swabbing RT-PCR-based surveillance collected across the country[1] (~300,000 swab tests per month; https://www.ons.gov.uk/peoplepopulationandcommunity/healthandsocialcare/conditionsanddiseases/methodologies/coronaviruscovid19infectionsurveyqmi for more details).

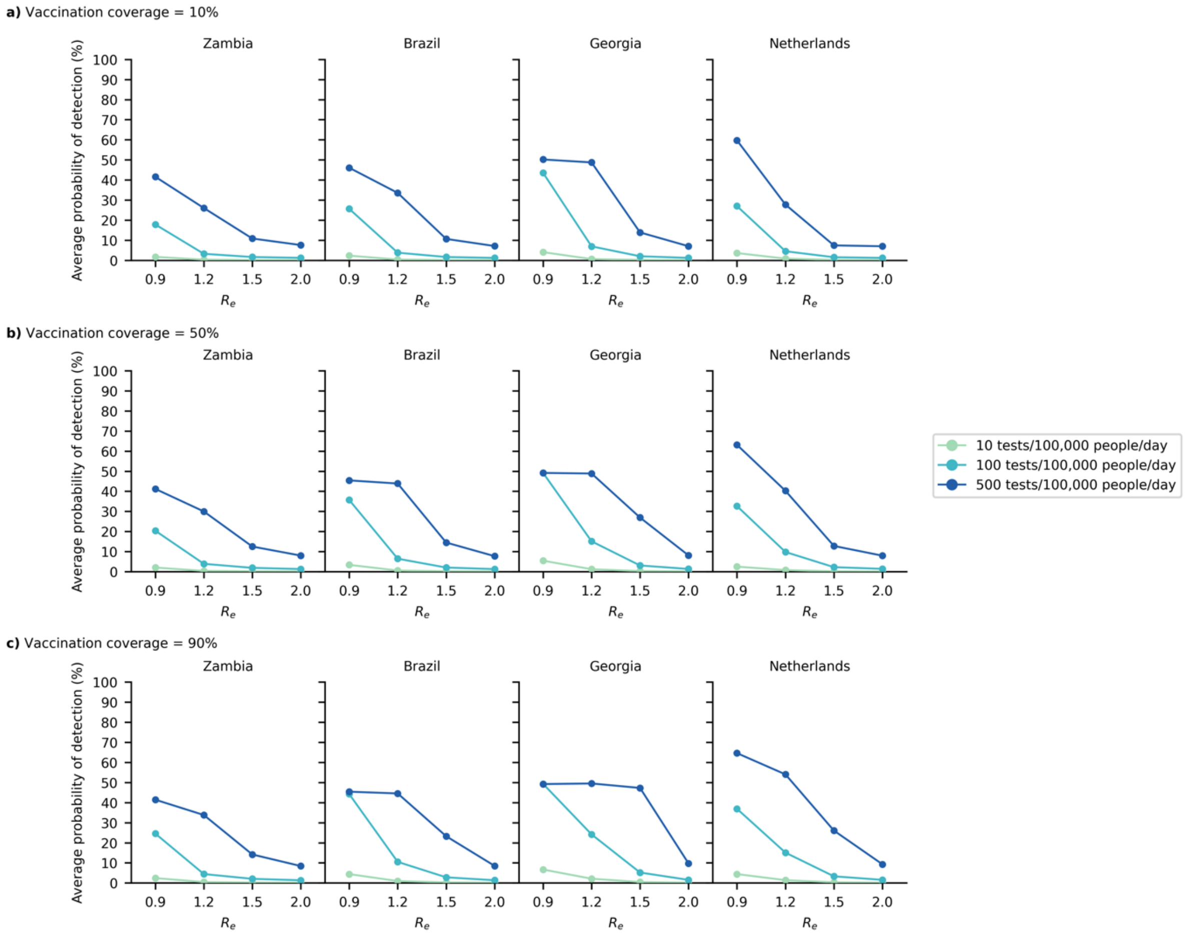


**Figure S2**: **Probability of detecting an infected case**. Line plots show the average effective probability that a symptomatic case would be detected in different countries (colours), under various epidemic intensity (i.e. $R_e$), test availability (colour shade), and vaccination coverage: (**a**) 10%, (**b**) 50% and (**c**) 90% for LMICs; 80% for the Netherlands. For Brazil, Georgia and Zambia (i.e. low-and-middle income countries (LMICs)), we assumed that tests are only available at health clinics and 65% of symptomatic individuals with mild disease would likely seek testing at clinics. For the Netherlands, we assumed that over-the-counter Ag-RDTs for self-testing are widely available (i.e. with no-cap on availability) such that only 10% of symptomatic individuals would seek clinic-provided testing directly. We also assumed that either 80% (solid line) or 50% (dashed line) of symptomatic individuals who did not seek clinic-provided testing would choose to perform a self-test using over-the-counter rapid tests instead.

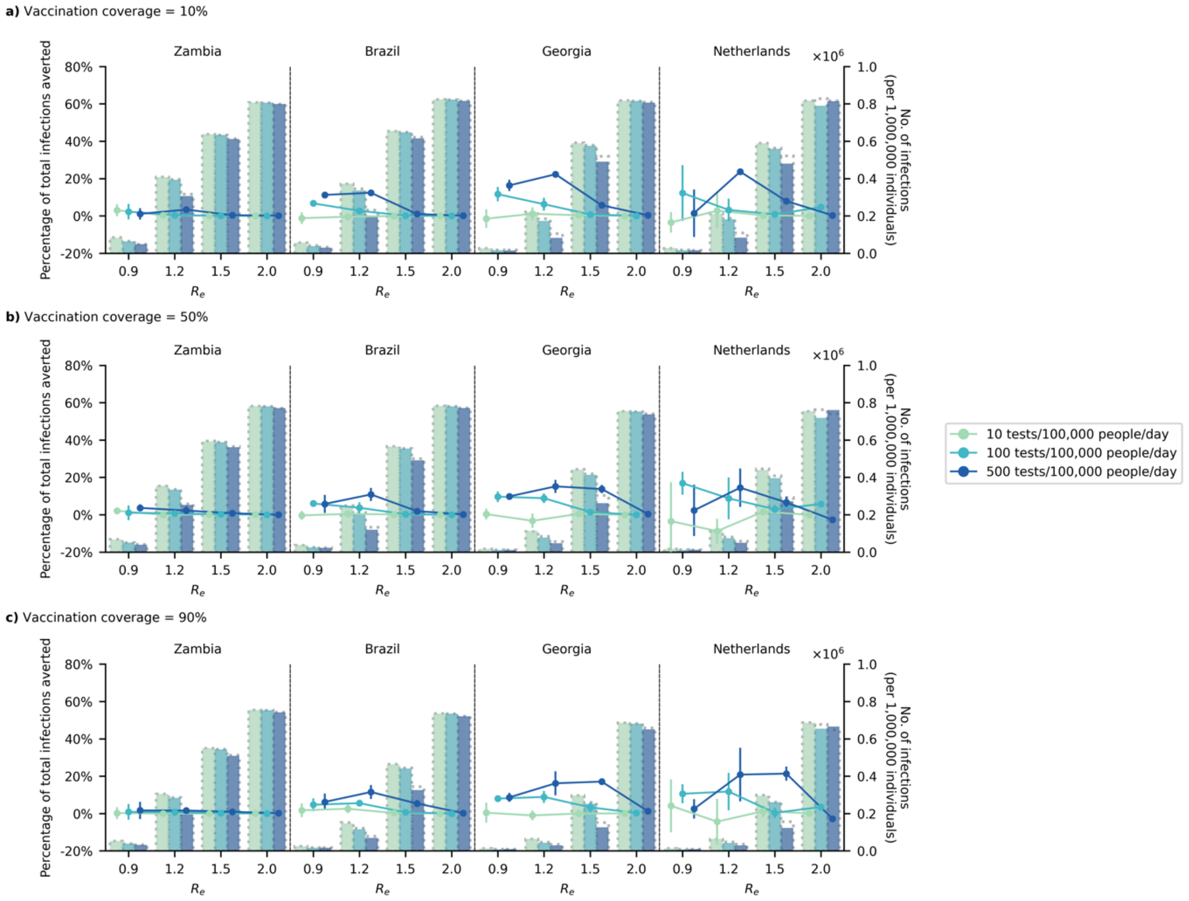


**Figure S3**: **Impact of test-and-treat on <u>infections</u>**. <u>No</u> restrictions on access to symptomatic testing at clinics (i.e. all symptomatic individuals who sought testing at clinics would receive one if in stock) and high-risk household contacts of test-positive individuals were <u>not</u> tested. All eligible high-risk individuals (i.e. ≥60 years of age or an adult ≥18 years with a relevant comorbidity) who tested positive were given a course of oral antivirals. Line plots (left $y$-axis) show the percentage change in total <u>infections</u> relative to no distribution of antivirals under different levels of mean test availability (different shades of color) after a 90-day epidemic wave in a population of 1,000,000 individuals with (**a**) 10%, (**b**) 50%, and (**c**) 90% vaccination coverage for different epidemic intensities (measured by the initial effective reproduction number ($R_e$); $x$-axis). Bar plots (right $y$-axis) show the number of <u>infections</u> in each corresponding scenario. The dotted outline of each bar shows the number of <u>infections</u> of each scenario if no antivirals were distributed.

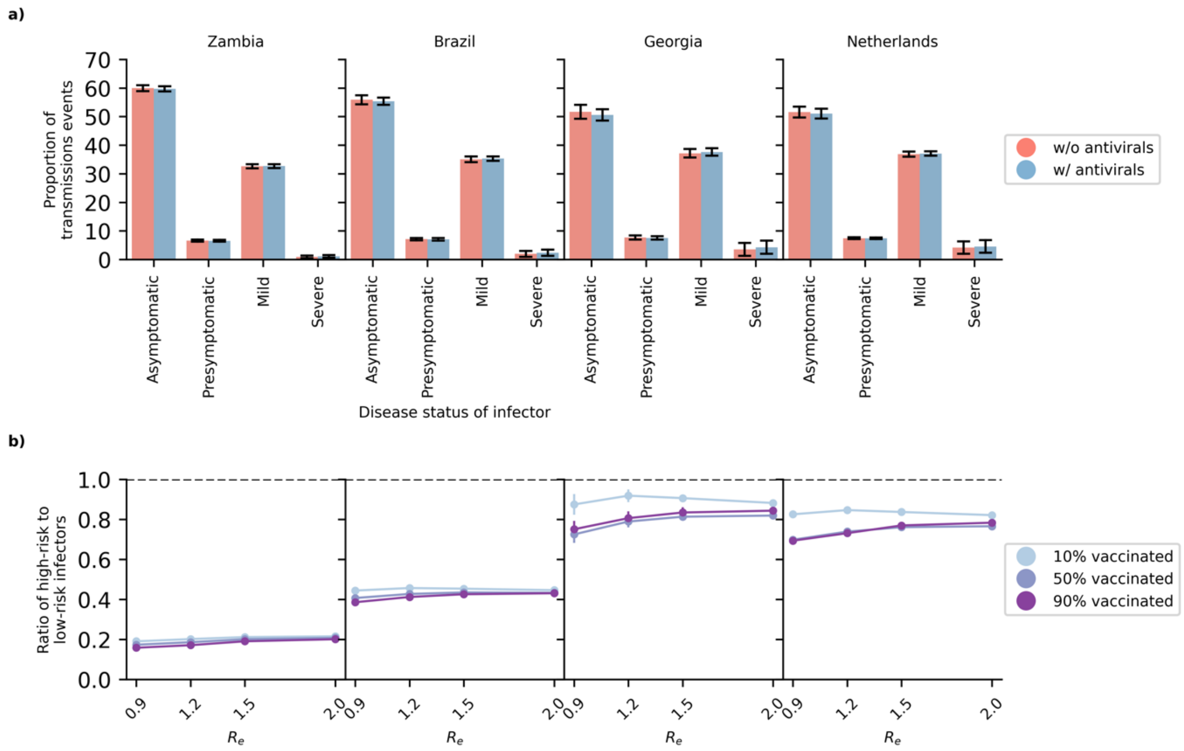


**Figure S4**: **Transmissions attributed to infectors of different disease and risk status**. (**a**) Bar plots show the mean proportion of transmissions events attributed to each type of infector (with standard deviation error bars), averaged across all simulated scenarios regardless if oral antivirals were distributed. (**b**) Line plots show the ratio of high-risk (i.e. ≥60 years of age or an adult ≥18 years with a relevant comorbidity) to low-risk infectors averaged across all test availability for different epidemic intensity (measured by effective reproduction number $R_e$).

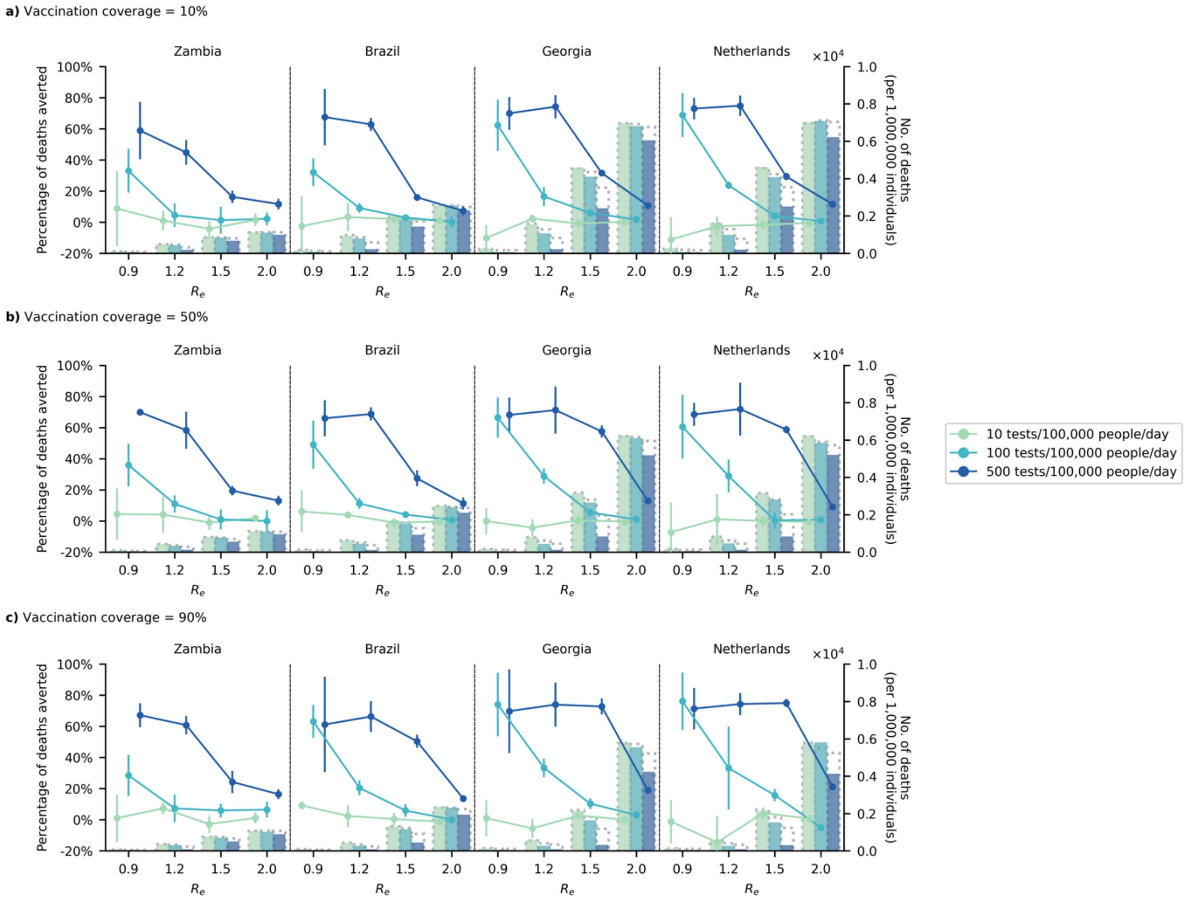


**Figure S5**: **Impact of test-and-treat on deaths**. No restrictions on access to symptomatic testing at clinics (i.e. all symptomatic individuals who sought testing at clinics would receive one if in stock) and high-risk household contacts of test-positive individuals were not tested. All eligible high-risk individuals (i.e. ≥60 years of age or an adult ≥18 years with a relevant comorbidity) who tested positive were given a course of oral antivirals. Line plots (left $y$-axis) show the percentage change in deaths relative to no distribution of antivirals under different levels of mean test availability (different shades of color) after a 90-day epidemic wave in a population of 1,000,000 individuals with (**a**) 10%, (**b**) 50%, and (**c**) 90% vaccination coverage for different epidemic intensities (measured by the initial effective reproduction number ($R_e$); $x$-axis). Bar plots (right $y$-axis) show the number of deaths in each corresponding scenario. The dotted outline of each bar shows the number of deaths of each scenario if no antivirals were distributed.

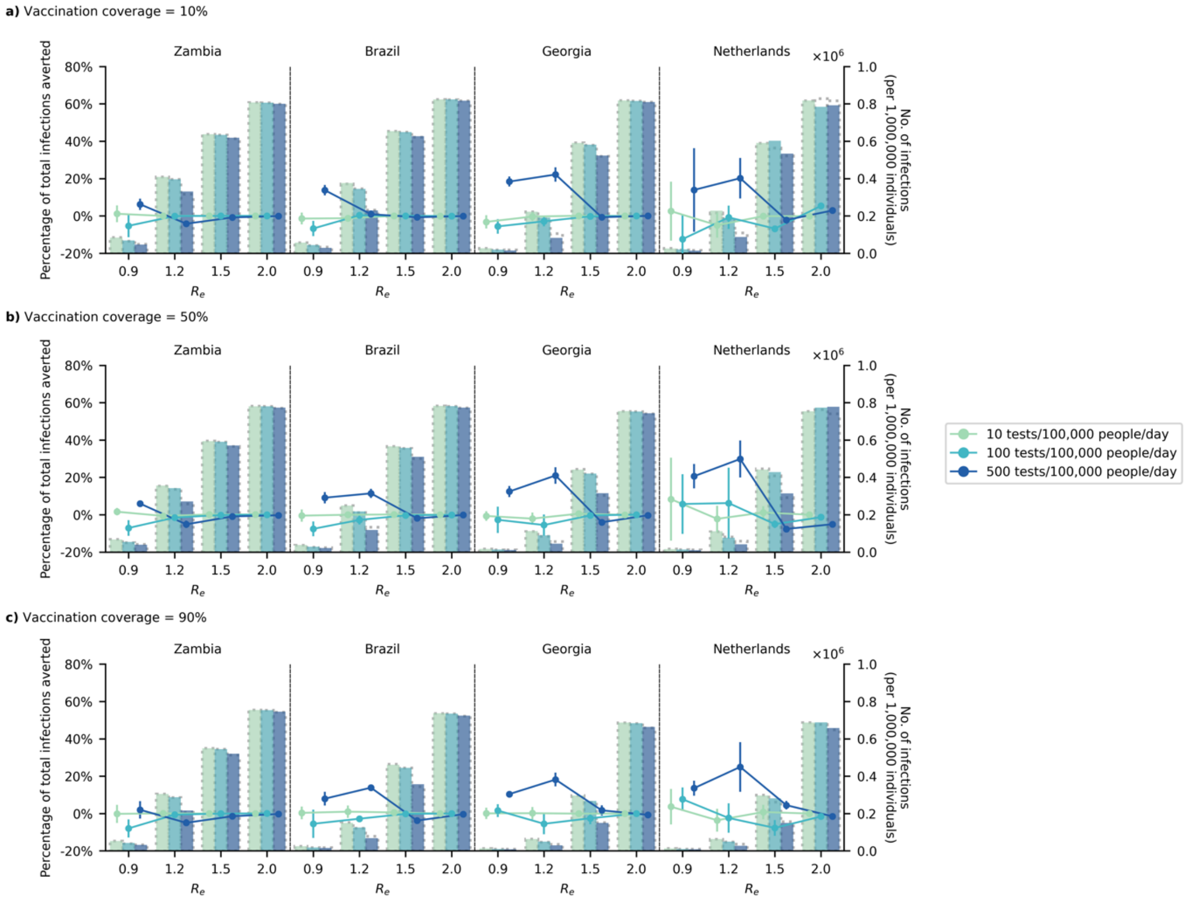


**Figure S6**: **Impact of test-and-treat with secondary distribution of test and antivirals to high-risk household contacts on infections**. No restrictions on access to symptomatic testing at clinics (i.e. all symptomatic individuals who sought testing at clinics would receive one if in stock). High-risk household contacts of test-positive individuals were given antigen rapid diagnostic tests to self-test for three consecutive days. All eligible high-risk individuals (i.e. ≥60 years of age or an adult ≥18 years with a relevant comorbidity) who tested positive, including high-risk household contacts who tested positive, were given a course of oral antivirals. Line plots (left $y$-axis) show the percentage change in total infections relative to no distribution of antivirals under different levels of mean test availability (different shades of color) after a 90-day epidemic wave in a population of 1,000,000 individuals with (**a**) 10%, (**b**) 50%, and (**c**) 90% vaccination coverage for different epidemic intensities (measured by the initial effective reproduction number ($R_e$); $x$-axis). Bar plots (right $y$-axis) show the number of infections in each corresponding scenario. The dotted outline of each bar shows the number of infections of each scenario if no antivirals were distributed.

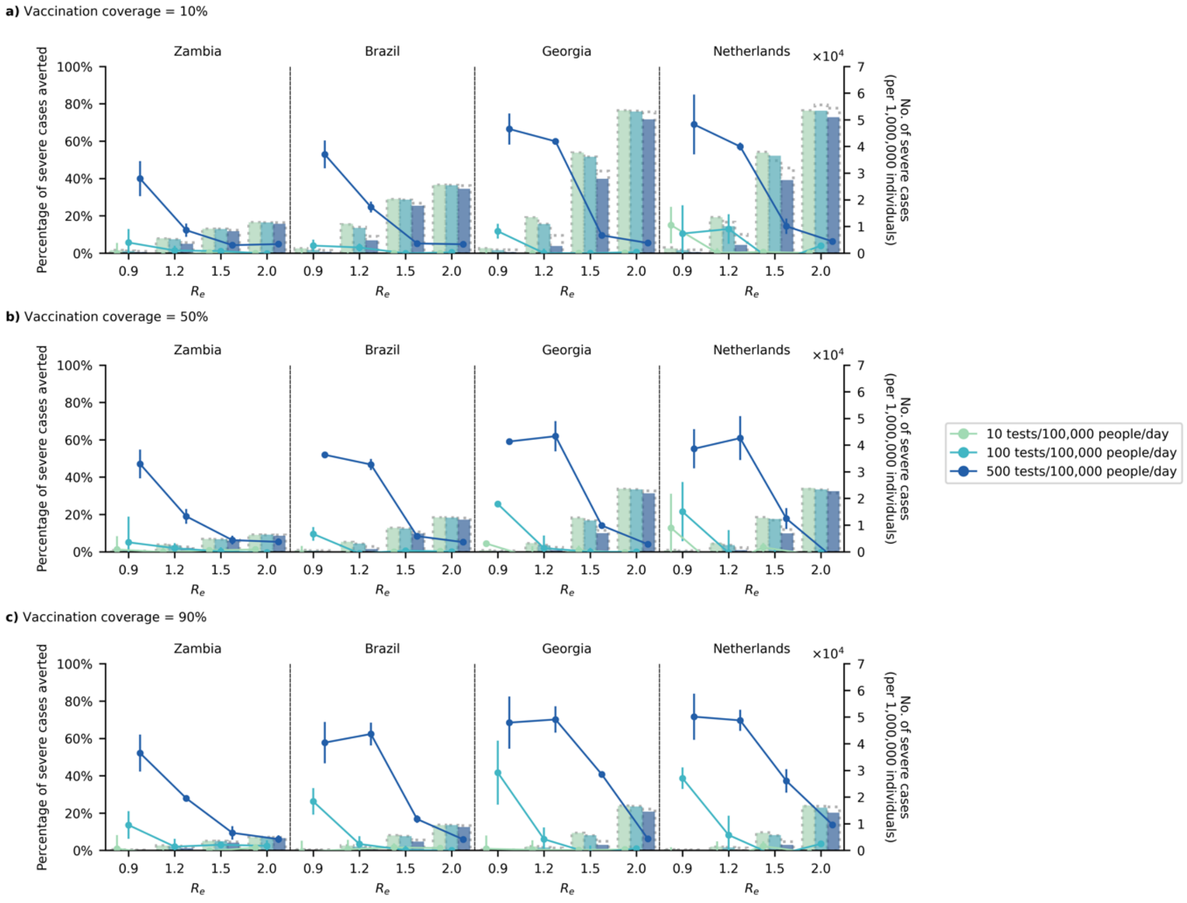


**Figure S7**: **Impact of test-and-treat with secondary distribution of test and antivirals to high-risk household contacts on severe cases**. No restrictions on access to symptomatic testing at clinics (i.e. all symptomatic individuals who sought testing at clinics would receive one if in stock). High-risk household contacts of test-positive individuals were given antigen rapid diagnostic tests to self-test for three consecutive days. All eligible high-risk individuals (i.e. ≥60 years of age or an adult ≥18 years with a relevant comorbidity) who tested positive, including high-risk household contacts who tested positive, were given a course of oral antivirals. Line plots (left $y$-axis) show the percentage in severe cases relative to no distribution of antivirals under different levels of mean test availability (different shades of color) after a 90-day epidemic wave in a population of 1,000,000 individuals with (**a**) 10%, (**b**) 50%, and (**c**) 90% vaccination coverage for different epidemic intensities (measured by the initial effective reproduction number ($R_e$); $x$-axis). Bar plots (right $y$-axis) show the number of infections in each corresponding scenario. The dotted outline of each bar shows the number of severe cases of each scenario if no antivirals were distributed.

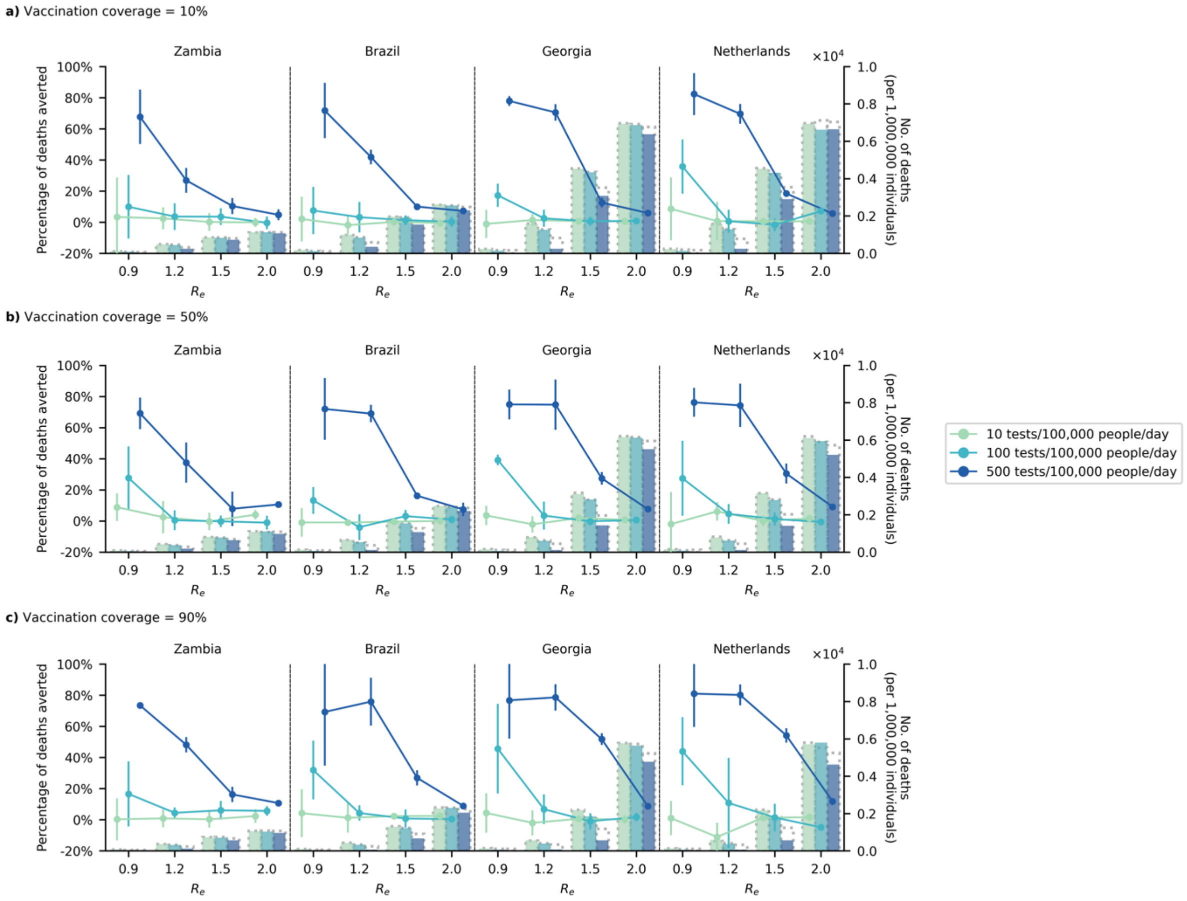


**Figure S8**: **Impact of test-and-treat with <u>secondary distribution of test and antivirals to high-risk household contacts</u> on <u>deaths</u>**. <u>No</u> restrictions on access to symptomatic testing at clinics (i.e. all symptomatic individuals who sought testing at clinics would receive one if in stock). <u>High-risk household contacts of test-positive individuals were given antigen rapid diagnostic tests to self-test for three consecutive days</u>. All eligible high-risk individuals (i.e. ≥60 years of age or an adult ≥18 years with a relevant comorbidity) who tested positive, <u>including high-risk household contacts who tested positive</u>, were given a course of oral antivirals. Line plots (left $y$-axis) show the percentage change in <u>deaths</u> relative to no distribution of antivirals under different levels of mean test availability (different shades of color) after a 90-day epidemic wave in a population of 1,000,000 individuals with (**a**) 10%, (**b**) 50%, and (**c**) 90% vaccination coverage for different epidemic intensities (measured by the initial effective reproduction number ($R_e$); $x$-axis). Bar plots (right $y$-axis) show the number of infections in each corresponding scenario. The dotted outline of each bar shows the number of <u>deaths</u> of each scenario if no antivirals were distributed.

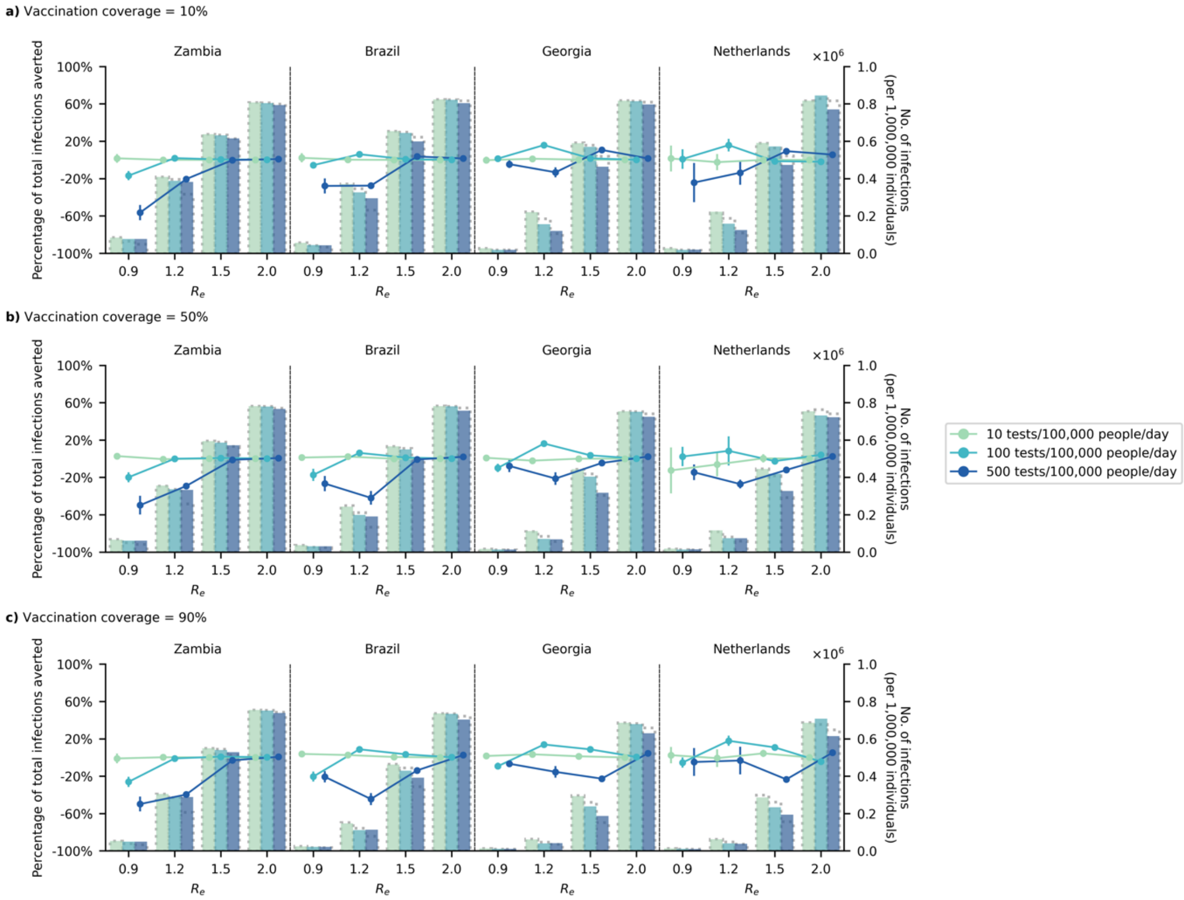


**Figure S9**: **Impact of test-and-treat on infections when restricting symptomatic testing to high-risk individuals only**. High-risk household contacts of test-positive individuals were not tested. All eligible high-risk individuals (i.e. ≥60 years of age or an adult ≥18 years with a relevant comorbidity) who tested positive were given a course of oral antivirals. Line plots (left $y$-axis) show the percentage change in total infections relative to no distribution of antivirals under different levels of mean test availability (different shades of color) after a 90-day epidemic wave in a population of 1,000,000 individuals with (**a**) 10%, (**b**) 50%, and (**c**) 90% vaccination coverage for different epidemic intensities (measured by the initial effective reproduction number ($R_e$); $x$-axis). Bar plots (right $y$-axis) show the number of infections in each corresponding scenario. The dotted outline of each bar shows the number of infections of each scenario if no antivirals were distributed.

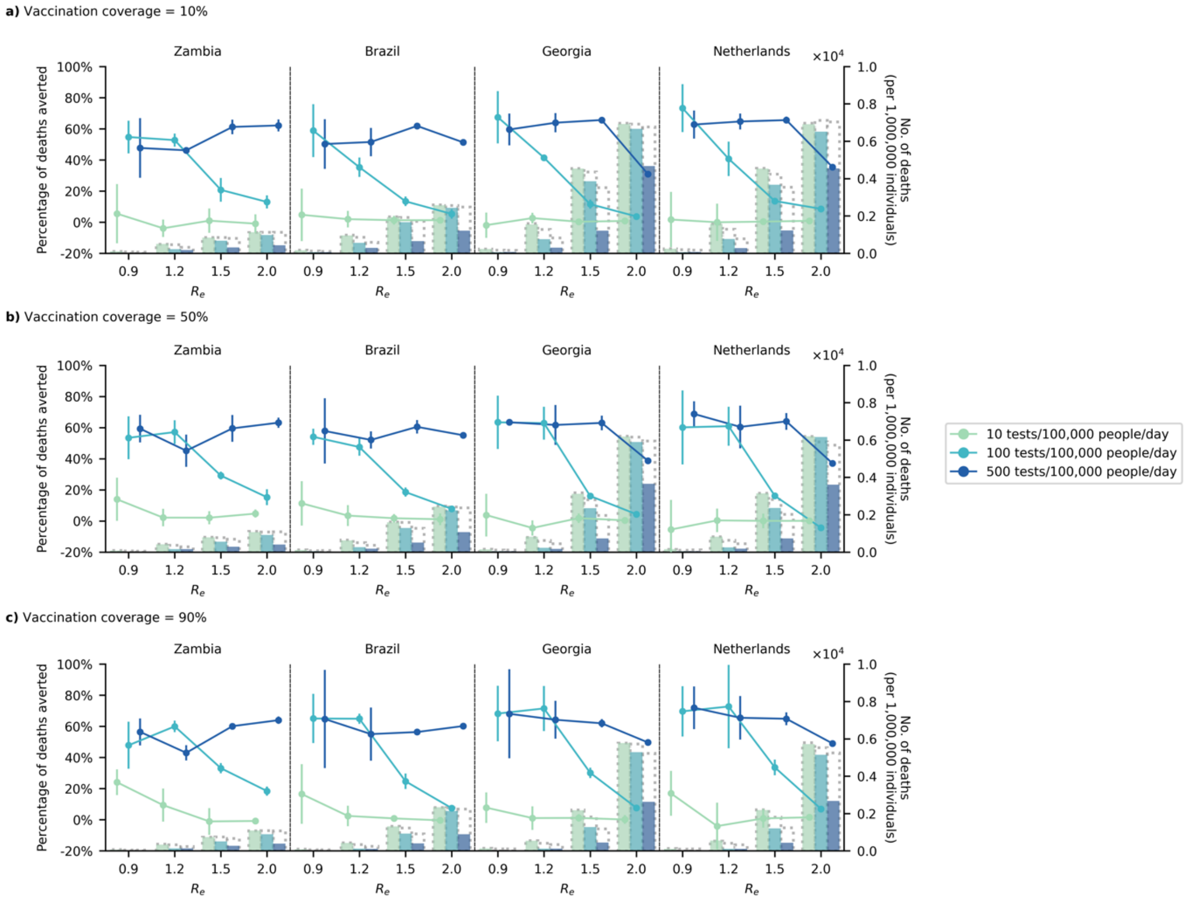


**Figure S10**: **Impact of test-and-treat on <u>deaths</u> when <u>restricting symptomatic testing at to high-risk individuals only</u>**. High-risk household contacts of test-positive individuals were <u>not</u> tested. All eligible high-risk individuals (i.e. ≥60 years of age or an adult ≥18 years with a relevant comorbidity) who tested positive were given a course of oral antivirals. Line plots (left $y$-axis) show the percentage change in <u>deaths</u> relative to no distribution of antivirals under different levels of mean test availability (different shades of color) after a 90-day epidemic wave in a population of 1,000,000 individuals with (**a**) 10%, (**b**) 50%, and (**c**) 90% vaccination coverage for different epidemic intensities (measured by the initial effective reproduction number ($R_e$); $x$-axis). Bar plots (right $y$-axis) show the number of <u>deaths</u> in each corresponding scenario. The dotted outline of each bar shows the number of <u>deaths</u> of each scenario if no antivirals were distributed.

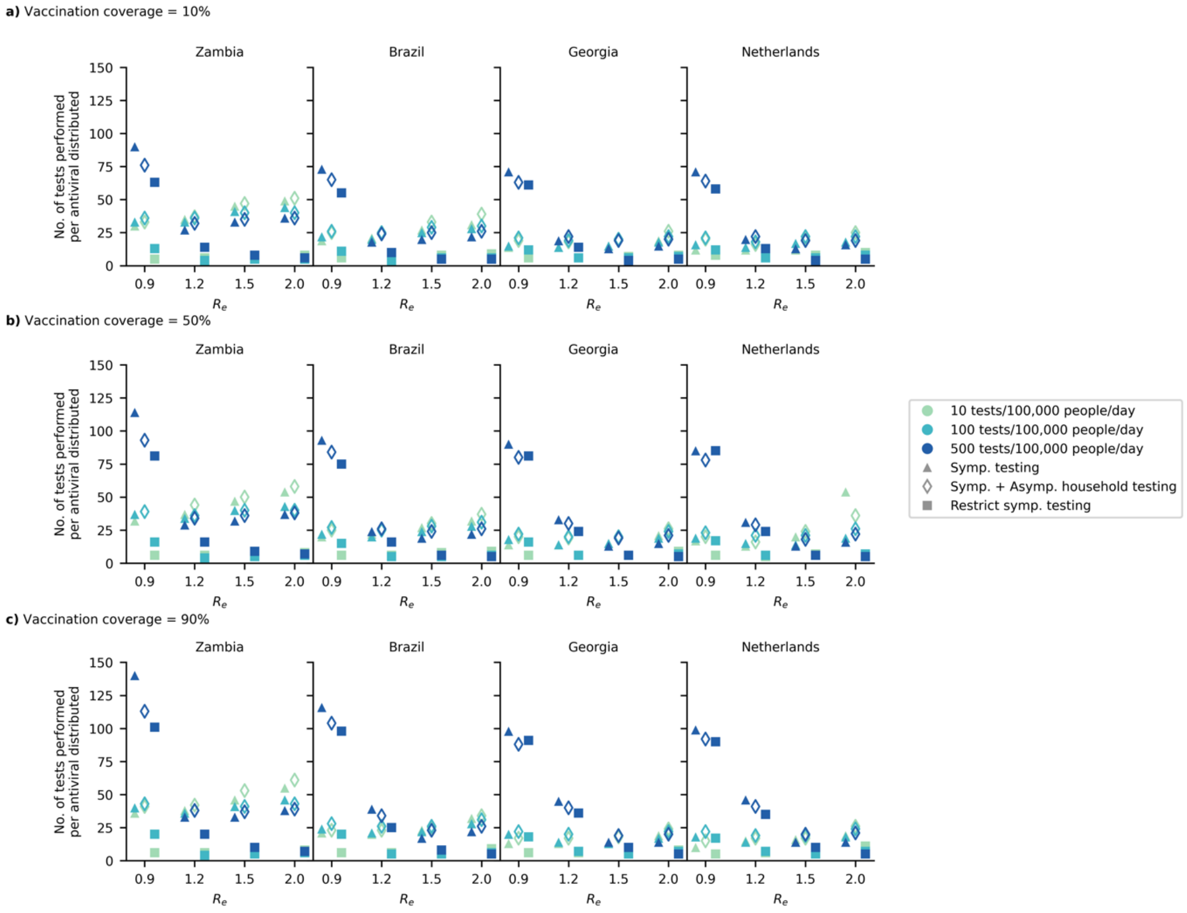


**Figure S11**: **Number of tests performed per antiviral distributed**. Each point shows the average number of tests performed per antiviral distributed under different testing rate (shading) in different countries (columns) under different testing strategy (triangles: only symptomatic persons who visit clinics will be tested if test is available; diamonds: asymptomatic household contacts of test-positive symptomatic person will also be tested over the next three consecutive days; squares: restrict symptomatic testing to high-risk individuals only). (**a**) 10%, (**b**) 50%, and (**c**) 90% vaccination coverage for different epidemic intensities (measured by the initial effective reproduction number ($R_e$); $x$-axis).

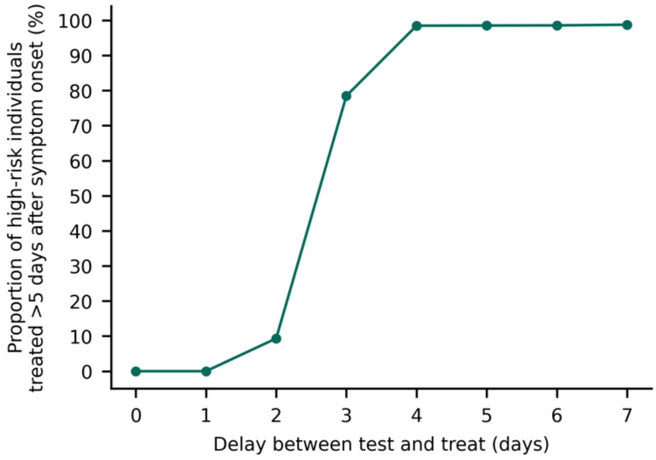


**Figure S12**: **Proportion of high-risk infected individuals treated >5 days after symptom onset if there were delays between test and treat**. Line plot shows the proportion of high-risk symptomatic individuals that would miss the treatment window of oral antivirals if they were tested at clinics *n* days late to receive a course of antiviral treatment.

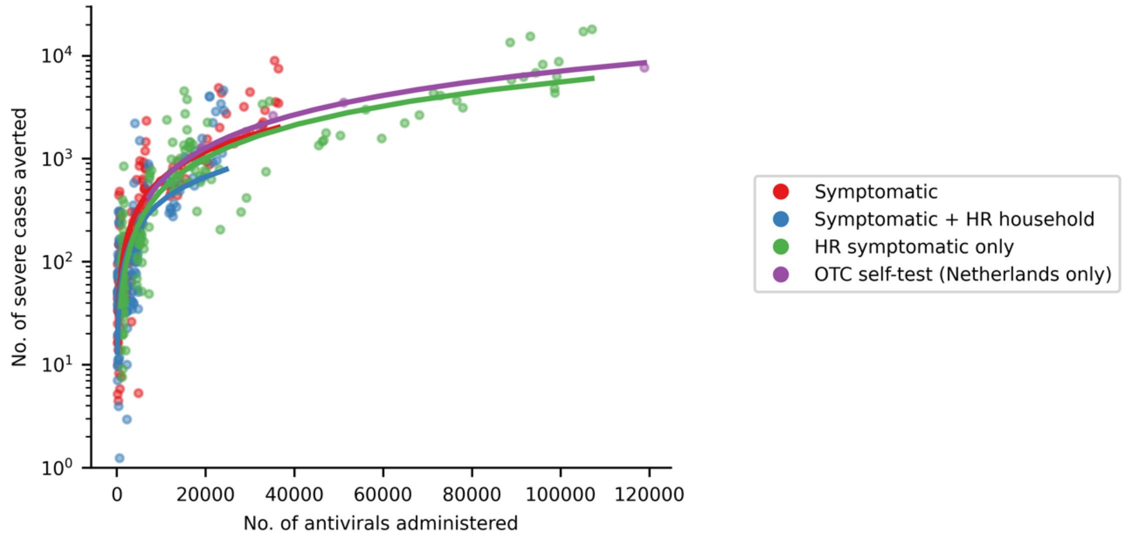


**Figure S13**: **Number of severe cases averted against number of oral antivirals administered for different test-and-treat strategies**. In all strategies, all eligible high-risk individuals (i.e. ≥60 years of age or an adult ≥18 years with a relevant comorbidity) who tested positive were given a course of oral antivirals. *Symptomatic* (Red): No restrictions on access to symptomatic testing at clinics (i.e. all symptomatic individuals who sought testing at clinics would receive one if in stock); *Symptomatic + HR household* (Blue): No restrictions on access to symptomatic testing at clinics. High-risk (HR) household contacts of test-positive individuals were given antigen rapid diagnostic tests to self-test for three consecutive days; *HR only* (Green): Symptomatic testing at clinics restricted to high-risk individuals only; *OTC self-test (Netherlands only)* (Purple): Wide availability of over-the-counter self-tests and large clinic-based test availability (i.e. 500 tests/100,000 people/day).

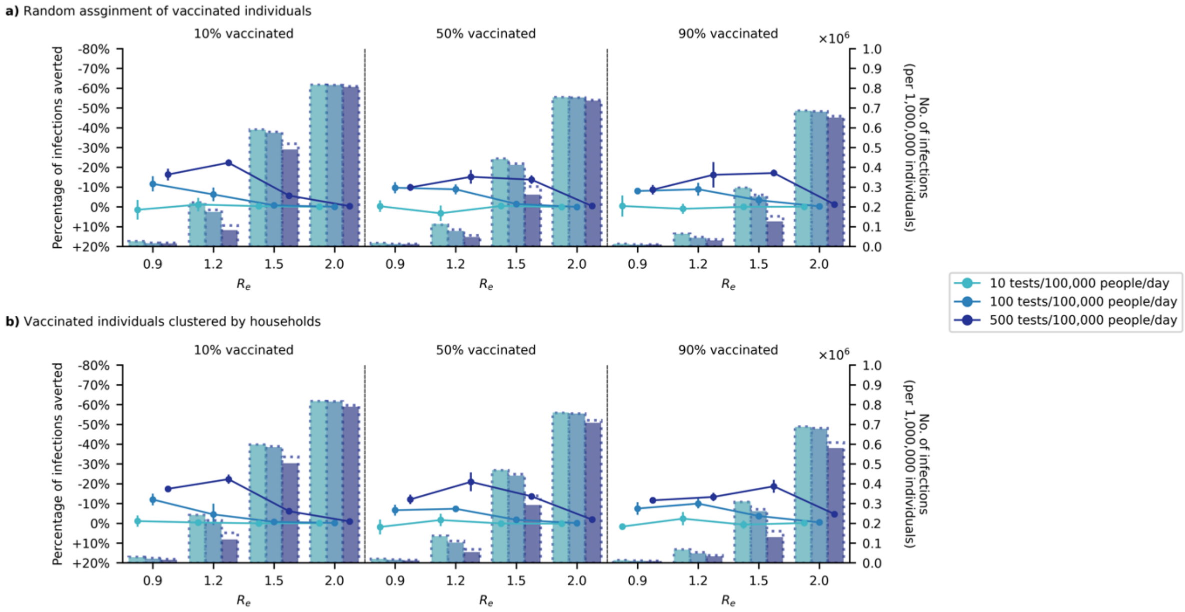


**Figure S14**: **Sensitivity analyses – Clustered vaccinated individuals; Impact of test-and-treat on infections in Georgia**. No restrictions on access to symptomatic testing at clinics (i.e. all symptomatic individuals who sought testing at clinics would receive one if in stock) and high-risk household contacts of test-positive individuals were not tested. All eligible high-risk individuals (i.e. ≥60 years of age or an adult ≥18 years with a relevant comorbidity) who tested positive were given a course of oral antivirals. Line plots (left $y$-axis) show the percentage change in total infections relative to no distribution of antivirals under different levels of mean test availability (different shades of color) after a 90-day epidemic wave in a population of 1,000,000 individuals with 10%, 50%, and 90% vaccination coverage for different epidemic intensities (measured by the initial effective reproduction number ($R_e$); $x$-axis). Bar plots (right $y$-axis) show the number of infections in each corresponding scenario. The dotted outline of each bar shows the number of infections of each scenario if no antivirals were distributed. (**a**) Random assignment of vaccinated individuals in the simulated population. (**b**) Vaccinated individuals are still randomly assigned but tend to be clustered by households.

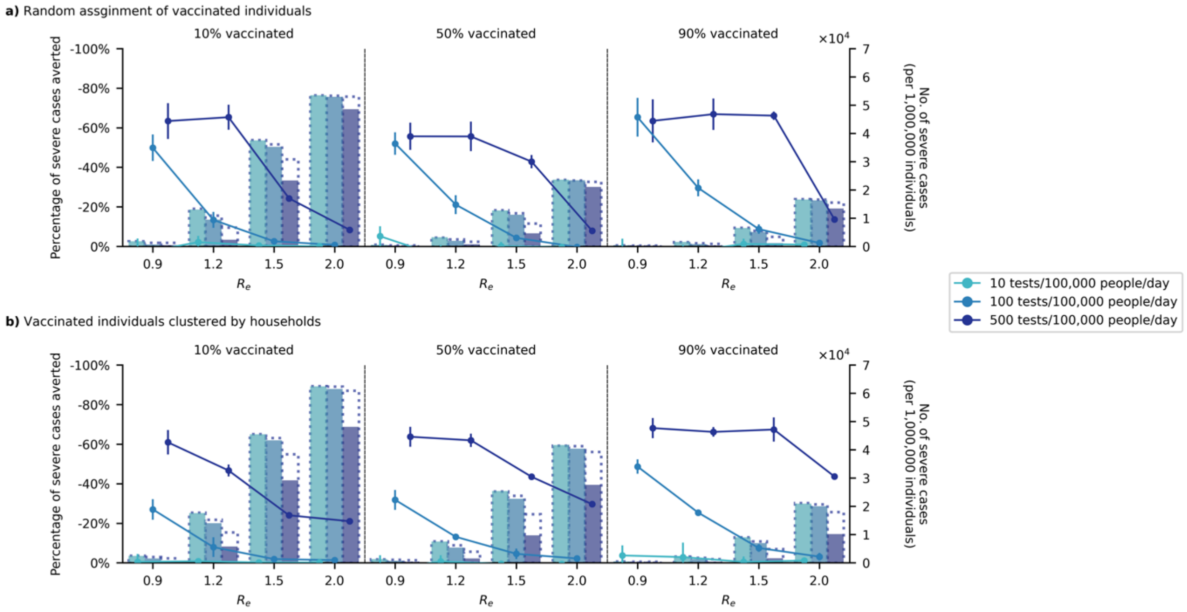


**Figure S15**: **Sensitivity analyses – Clustered vaccinated individuals; Impact of test-and-treat on severe cases in Georgia**. No restrictions on access to symptomatic testing at clinics (i.e. all symptomatic individuals who sought testing at clinics would receive one if in stock) and high-risk household contacts of test-positive individuals are not tested. All eligible high-risk individuals (i.e. ≥60 years of age or an adult ≥18 years with a relevant comorbidity) who tested positive were given a course of oral antivirals. Line plots (left $y$-axis) show the percentage change in severe cases relative to no distribution of antivirals under different levels of mean test availability (different shades of color) after a 90-day epidemic wave in a population of 1,000,000 individuals with (a) 10%, (b) 50%, and (c) 90% vaccination coverage for different epidemic intensities (measured by the initial effective reproduction number ($R_e$); $x$-axis). Bar plots (right $y$-axis) show the number of severe cases in each corresponding scenario. The dotted outline of each bar shows the number of severe cases of each scenario when no antivirals were distributed. (**a**) Random assignment of vaccinated individuals in the simulated population. (**b**) Vaccinated individuals are still randomly assigned but tend to be clustered by households.

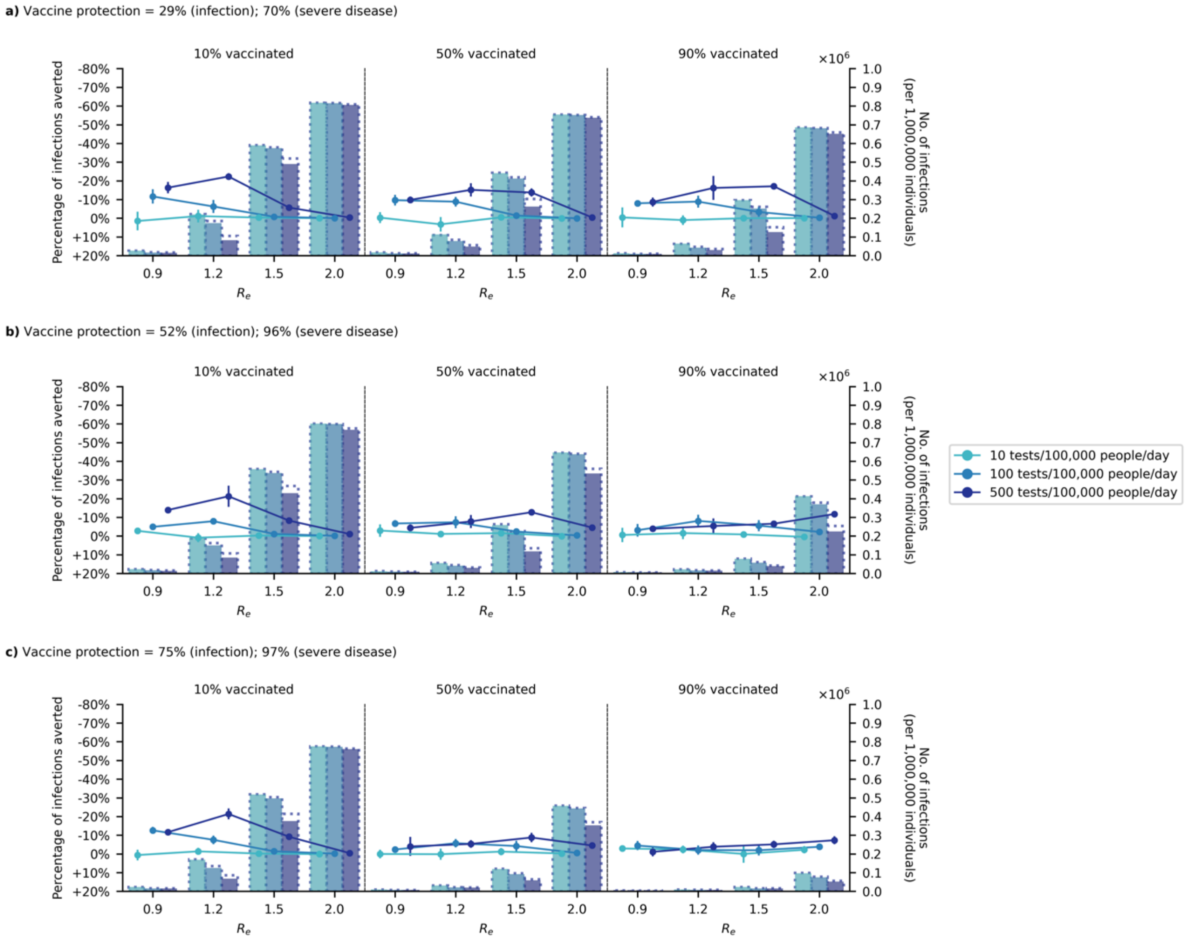


**Figure S16**: **Sensitivity analyses – Varying vaccine effectiveness; Impact of test-and-treat on infections in Georgia**. No restrictions on access to symptomatic testing at clinics (i.e. all symptomatic individuals who sought testing at clinics would receive one if in stock) and high-risk household contacts of test-positive individuals were not tested. All eligible high-risk individuals (i.e. ≥60 years of age or an adult ≥18 years with a relevant comorbidity) who tested positive were given a course of oral antivirals. Line plots (left $y$-axis) show the percentage change in total infections relative to no distribution of antivirals under different levels of mean test availability (different shades of color) after a 90-day epidemic wave in a population of 1,000,000 individuals with 10%, 50%, and 90% vaccination coverage for different epidemic intensities (measured by the initial effective reproduction number ($R_e$); $x$-axis). Bar plots (right $y$-axis) show the number of infections in each corresponding scenario. The dotted outline of each bar shows the number of infections of each scenario if no antivirals were distributed. (**a**) 29%/70% (**b**) 52%/96% and (**c**) 75%/97% protection against infection/severe disease.

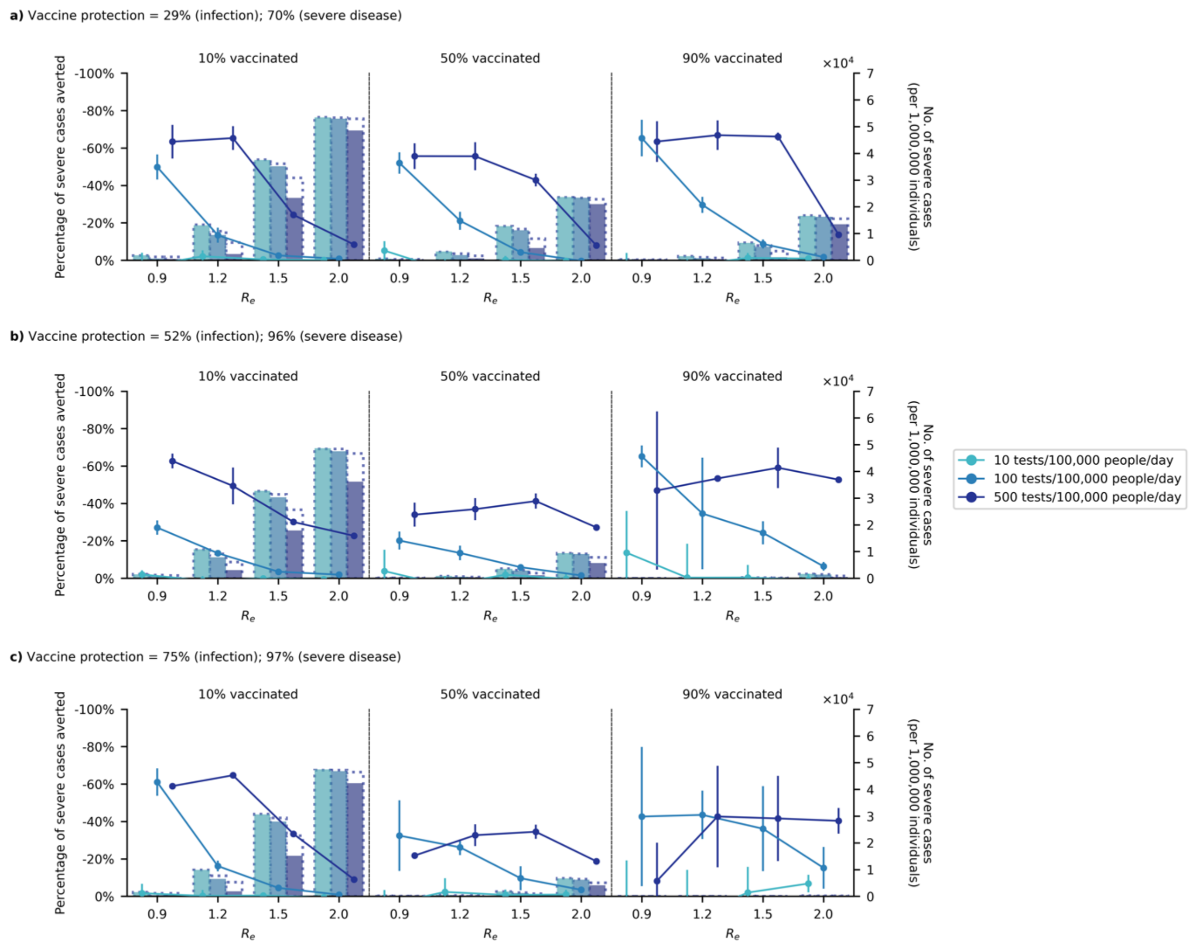


**Figure S17**: **Sensitivity analyses – Varying vaccine effectiveness; Impact of test-and-treat on severe cases in Georgia**. No restrictions on access to symptomatic testing at clinics (i.e. all symptomatic individuals who sought testing at clinics would receive one if in stock) and high-risk household contacts of test-positive individuals are not tested. All eligible high-risk individuals (i.e. ≥60 years of age or an adult ≥18 years with a relevant comorbidity) who tested positive were given a course of oral antivirals. Line plots (left $y$-axis) show the percentage change in severe cases relative to no distribution of antivirals under different levels of mean test availability (different shades of color) after a 90-day epidemic wave in a population of 1,000,000 individuals with (a) 10%, (b) 50%, and (c) 90% vaccination coverage for different epidemic intensities (measured by the initial effective reproduction number ($R_e$); $x$-axis). Bar plots (right $y$-axis) show the number of severe cases in each corresponding scenario. The dotted outline of each bar shows the number of severe cases of each scenario when no antivirals were distributed. (**a**) 29%/70% (**b**) 52%/96% and (**c**) 75%/97% protection against infection/severe disease.

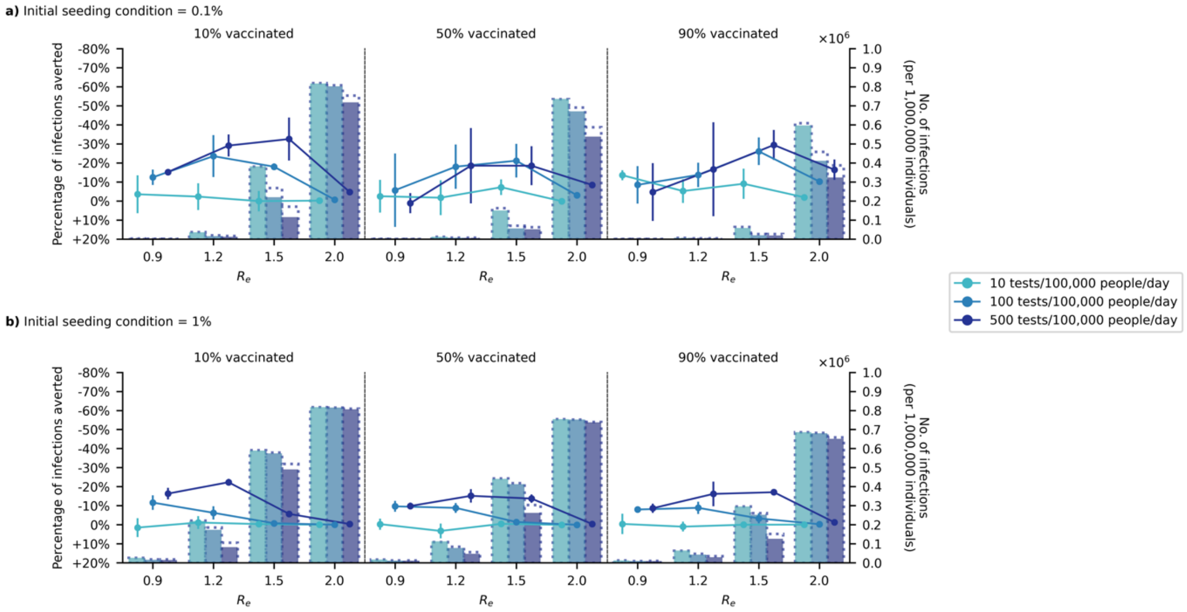


**Figure S18**: **Sensitivity analyses – Lower seeding condition; Impact of test-and-treat on <u>infections</u> in Georgia**. <u>No</u> restrictions on access to symptomatic testing at clinics (i.e. all symptomatic individuals who sought testing at clinics would receive one if in stock) and high-risk household contacts of test-positive individuals were <u>not</u> tested. All eligible high-risk individuals (i.e. ≥60 years of age or an adult ≥18 years with a relevant comorbidity) who tested positive were given a course of oral antivirals. Line plots (left $y$-axis) show the percentage change in total <u>infections</u> relative to no distribution of antivirals under different levels of mean test availability (different shades of color) after a 90-day epidemic wave in a population of 1,000,000 individuals with 10%, 50%, and 90% vaccination coverage for different epidemic intensities (measured by the initial effective reproduction number ($R_e$); $x$-axis). Bar plots (right $y$-axis) show the number of <u>infections</u> in each corresponding scenario. The dotted outline of each bar shows the number of <u>infections</u> of each scenario if no antivirals were distributed. Initial proportion of population infected: (**a**) 0.1% (**b**) 1%.

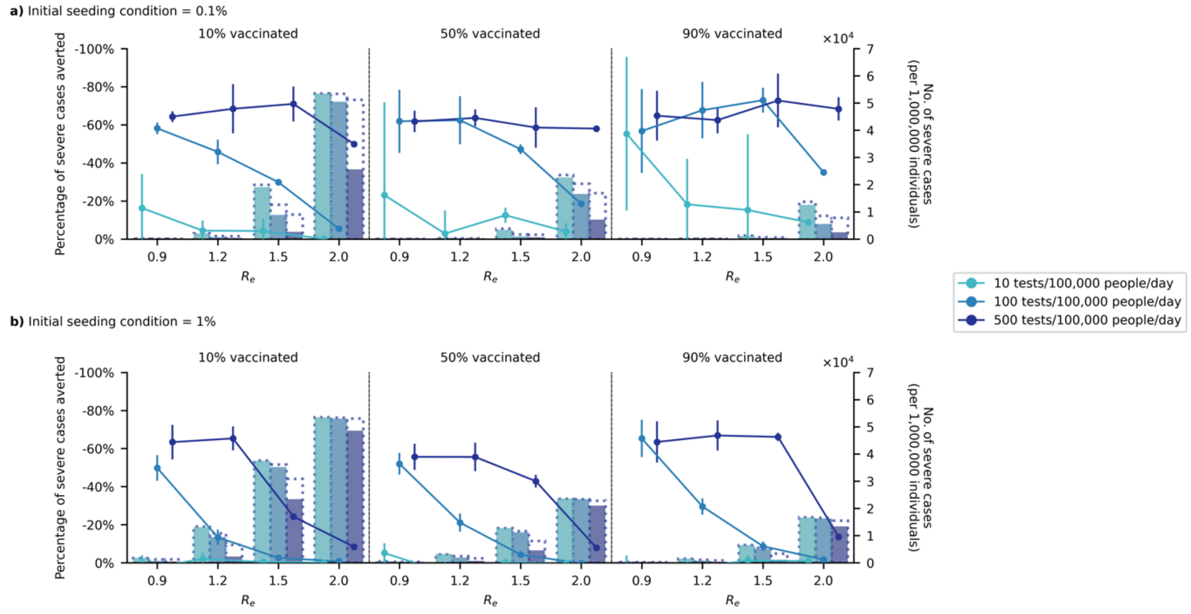


**Figure S19**: **Sensitivity analyses – Lower seeding condition; Impact of test-and-treat on severe cases in Georgia**. No restrictions on access to symptomatic testing at clinics (i.e. all symptomatic individuals who sought testing at clinics would receive one if in stock) and high-risk household contacts of test-positive individuals are not tested. All eligible high-risk individuals (i.e. ≥60 years of age or an adult ≥18 years with a relevant comorbidity) who tested positive were given a course of oral antivirals. Line plots (left $y$-axis) show the percentage change in severe cases relative to no distribution of antivirals under different levels of mean test availability (different shades of color) after a 90-day epidemic wave in a population of 1,000,000 individuals with (a) 10%, (b) 50%, and (c) 90% vaccination coverage for different epidemic intensities (measured by the initial effective reproduction number ($R_e$); $x$-axis). Bar plots (right $y$-axis) show the number of severe cases in each corresponding scenario. The dotted outline of each bar shows the number of severe cases of each scenario when no antivirals were distributed. Initial proportion of population infected: (**a**) 0.1% (**b**) 1%.

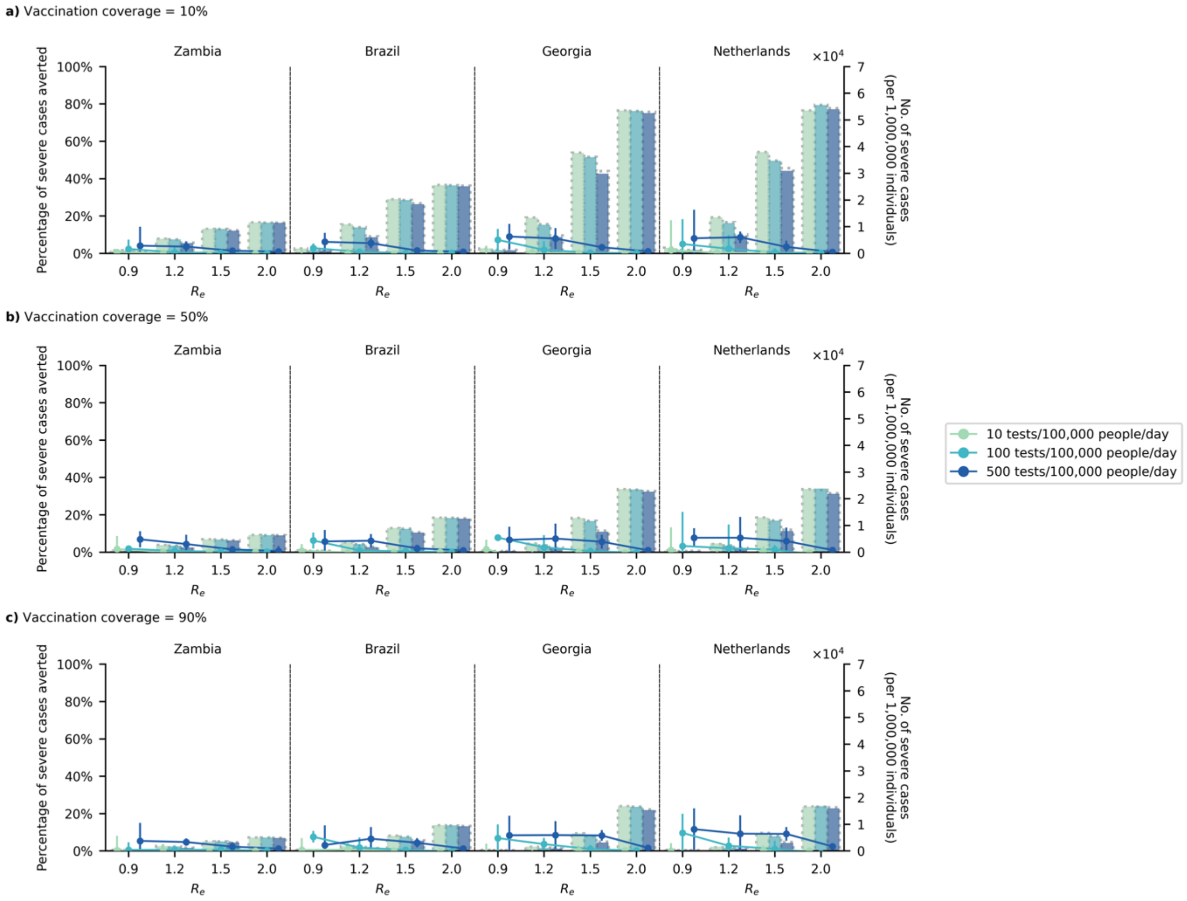


**Figure S20: Sensitivity analyses – Lower willingness to test; Impact of test-and-treat on severe cases**. All other simulations have assumed there is a 65% chance a symptomatic individual would seek testing. Here, we assumed that this probability has dwindled to 10% only. No restrictions on access to symptomatic testing at clinics (i.e. all symptomatic individuals who sought testing at clinics would receive one if in stock) and high-risk household contacts of test-positive individuals are not tested. All eligible high-risk individuals (i.e. ≥60 years of age or an adult ≥18 years with a relevant comorbidity) who tested positive were given a course of oral antivirals. Line plots (left $\boldsymbol{y}$-axis) show the percentage change in severe cases relative to no distribution of antivirals under different levels of mean test availability (different shades of color) after a 90-day epidemic wave in a population of 1,000,000 individuals with (a) 10%, (b) 50%, and (c) 90% vaccination coverage for different epidemic intensities (measured by the initial effective reproduction number ($\boldsymbol{R_e}$); $\boldsymbol{x}$-axis). Bar plots (right $\boldsymbol{y}$-axis) show the number of severe cases in each corresponding scenario. The dotted outline of each bar shows the number of severe cases of each scenario when no antivirals were distributed.

#### Supplementary Tables

#### Table S1: Variables and parameters used in PATAT.

| Parameter | Values/Distribution | Source |
|---|---|---|
| *Population demography* | | |
| Total population size | 1,000,000 | |
| Mean household size | Zambia: 5.0<br>Brazil, Georgia: 3.3<br>Netherlands: 2.1 | [2–5] |
| Age structure (in bins of 5 years) | Zambia: [0.161, 0.165, 0.157, 0.101, 0.083, 0.068, 0.057, 0.051, 0.042, 0.030, 0.024, 0.015, 0.016, 0.009, 0.008, 0.005, 0.006, 0.002, 0.000, 0.000]<br>Brazil: [0.072, 0.078, 0.090, 0.089, 0.090, 0.090, 0.083, 0.073, 0.068, 0.062, 0.053, 0.043, 0.034, 0.025, 0.020, 0.013, 0.009, 0.004, 0.002, 0.001]<br>Georgia: [0.013, 0.057, 0.071, 0.064, 0.055, 0.058, 0.065, 0.073, 0.070, 0.065, 0.063, 0.062, 0.069, 0.064, 0.053, 0.039, 0.024, 0.025, 0.006, 0.006]<br>Netherlands: [0.049, 0.051, 0.054, 0.058, 0.064, 0.064, 0.065, 0.061, 0.059, 0.062, 0.073, 0.072, 0.066, 0.058, 0.054, 0.041, 0.026, 0.015, 0.006, 0.002] | [2,3,5,6] |
| Minimum prime adult age | 20 years | Assumed |
| Proportion of women | 51% (Zambia, Brazil), 52% (Georgia), 50% (Netherlands) | [5–8] |
| Minimum working age | 15 years (Zambia, Brazil, Georgia), 16 years (Netherlands) | [5–8] |
| Employment rate | Zambia: 39% (male), 23% (female)<br>Brazil: 73% (male), 53% (female)<br>Georgia: 77% (male), 82% (female)<br>Netherlands: 75% (male), 68% (female) | [5–8] |
| Formal employment rate | Zambia: 36% (male), 24% (female)<br>Brazil: 90% (male), 90% (female)<br>Georgia: 64% (male), 74% (female)<br>Netherlands: 81% (male), 88% (female) | [5–8] |
| Schooling rate | Zambia: 79% (Primary), 40% (Secondary)<br>Brazil: 97% (Primary), 83% (Secondary)<br>Georgia: 98% (Primary), 95% (Secondary)<br>Netherlands: 99% (Primary), 92% (Secondary) | [2,3,9,10] |
| School gender parity | Zambia: 1.00 (Primary), 0.90 (Secondary)<br>Brazil: 0.97 (Primary), 0.98 (Secondary)<br>Georgia: 1.00 (Primary and secondary)<br>Netherlands: 1.00 (Primary and secondary) | [2,3,9,10] |
| Religious gathering participation rate | Zambia: 70%<br>Brazil: 41%<br>Georgia: 13% | [11] |

| | | |
|---|---|---|
| | Netherlands: NA (Assumed) | |
| Mean employment contacts (formal) | 20 | Assumed |
| Mean employment contacts (informal) | 5 | Assumed |
| Mean class size | Zambia: 37 (Primary and secondary)<br>Brazil: 20 (Primary), 26 (Secondary)<br>Georgia: 20 (Primary and Secondary)<br>Netherlands: 25 (Primary and secondary, assumed) | [2,12,13] |
| Mean school size | Zambia: 700 (Primary and secondary, assumed)<br>Brazil: 500 (Primary), 400 (Secondary) (Assumed)<br>Georgia: 135 (Primary and secondary)<br>Netherlands: 224 (Primary), 1442 (Secondary) | [5,6] |
| Student-teacher ratio | Zambia: 42 (Primary and secondary)<br>Brazil: 20 (Primary), 17 (Secondary)<br>Georgia: 8 (Primary and secondary)<br>Netherlands: 12 (Primary), 15 (Secondary) | [2,14] |
| Mean religious gathering size (s.d.) | Zambia: 500 (100)<br>Brazil, Georgia: 200 (100)<br>Netherlands: NA | Assumed |
| Mean random contacts in religious gathering per person | 10 | Assumed |
| Mean random community contacts per day | 10 | Assumed |
| *SARS-CoV-2 transmissions related parameters* | | |
| Age-structured relative susceptibility (in bins of 5 years) | [0.34, 0.34, 0.67, 0.67, 1.00, 1.00, 1.00, 1.00, 1.00, 1.00, 1.00, 1.00, 1.00, 1.00, 1.24, 1.24, 1.47, 1.47, 1.47, 1.47] | [15,16] |
| Age-structured probability of becoming symptomatic (in bins of 5 years) | [0.50, 0.50, 0.55, 0.55, 0.60, 0.60, 0.65, 0.65, 0.70, 0.70, 0.75, 0.75, 0.80, 0.80, 0.85, 0.85, 0.90, 0.90, 0.90, 0.90] | [17,18] |
| Age-structured probability of developing severe disease (in bins of 5 years) | [0.00050, 0.00050, 0.00165, 0.00165, 0.00720, 0.00720, 0.02080, 0.02080, 0.03430, 0.03430, 0.07650, 0.07650, 0.13280, 0.13280, 0.20655, 0.20655, 0.24570, 0.24570, 0.24570, 0.24570] | [17,18] |
| Age-structured probability of death (in bins of 5 years) | [0.00002, 0.00002, 0.00002, 0.00002, 0.00010, 0.00010, 0.00032, 0.00032, 0.00098, 0.00098, 0.00265, 0.00265, 0.00766, 0.00766, 0.02439, 0.02439, 0.08292, 0.08292, 0.16190, 0.16190] | [19,20] |
| Latent period (days) | Lognormal; Mean = 4.0, SD = 1.3 | [16,21–23] |
| Pre-symptomatic period (days) | Lognormal; Mean = 1.8, SD = 1.7 | [16,21,23] |
| Period between symptom onset and severe disease (days) | Lognormal; Mean = 6.6, SD = 4.9 | [21] |
| Period between severe disease and death (days) | Lognormal; Mean = 8.6, SD = 6.7 | [21] |
| Recovery period for symptomatic agents with mild disease (days) | Lognormal; Mean = 5.35, SD = 0.37[*] | [23,24] |
| Recovery period for asymptomatic agent (days) | Lognormal; Mean = 5.35, SD = 0.37[*] | [23,24] |

| | | |
|---|---|---|
| Recovery period of agents with severe disease (days) | Lognormal; Mean = 18.1, SD = 6.3 | 17 |
| Peak Ct values | Normal; Mean = 23.3, SD = 0.58* | 23 |
| Cross-immunity to variant virus after infection by extant virus | 20% | 25,26 |
| Severity (chance of hospitalization) of variant relative to extant virus | 40% | 27 |
| *Testing parameters* | | |
| Delay in visiting healthcare facility for symptomatic testing (days) | Lognormal; Mean = 1.0, SD = 0.5 | Assumed |
| Ag-RDT specificity | 0.989 | 28 |
| *Isolation/quarantine parameters* | | |
| Isolation period | 10 days | |
| Quarantine period | 14 days | |
| Self-isolation period | 10 days | |
| Reduction in contact rates under isolation/quarantine (in order of households, schools, workplaces, religious gathering and random community) | [10%, 100%, 100%, 100%, 100%] | |

*Standard deviation values inferred from 95% confidence interval computed in reference.
